# Prevalence and characteristics of *Plasmodium vivax* Gametocytes in Duffy-positive and Duffy-negative populations across Ethiopia

**DOI:** 10.1101/2023.12.10.23299780

**Authors:** Ebony Little, Tassew T. Shenkutie, Meshesha Tsigie Negash, Beka R. Abagero, Abnet Abebe, Jean Popovici, Sindew Mekasha, Eugenia Lo

## Abstract

*Plasmodium* parasites replicate asexually in the human host. The proportion of infections that carries gametocytes is a proxy for human-to-mosquito transmissibility. It is unclear what proportion of *P. vivax* infections in Duffy-negatives carries gametocytes. This study aims to determine the prevalence of *P. vivax* in Duffy-negatives across broad regions of Ethiopia and characterize parasite stages. Finger-prick blood samples were collected for microscopic and molecular screening of *Plasmodium* parasites and Duffy status of individuals. Molecular screening of plasmodium species and Duffy blood group genotyping was done using SYBR green and Taqman qPCR method. Among the total 447 samples, 414 (92.6%) were *P*. *vivax* confirmed and, 16 (3.9%) of them were from Duffy-negatives. Of these, 5/16 (31.3%) Duffy-negative *P*. *vivax*-infected samples were detected with gametocytes. Of the 398 Duffy-positive *P*. *vivax*-infected samples, 150 (37.7%) were detected with gametocytes, slightly higher than that in Duffy-negatives. This study highlights the presence of *P. vivax* gametocytes in Duffy-negative infections, suggestive of human-to-mosquito transmissibility. Although *P. vivax* infections in Duffy-negatives are commonly associated with low parasitemia, some of these infections were shown with relatively high parasitemia and may represent better erythrocyte invasion capability of *P. vivax* and hidden reservoirs that can contribute to transmission. A better understanding of *P. vivax* transmission biology and gametocyte function particularly in Duffy-negative populations would aid future treatment and management of vivax malaria in Africa

## Introduction

Yearly, there are about 619,000 malaria-related deaths and ∼247 million malaria cases reported globally.^1^ Out of the five malaria *Plasmodium* species, *P. vivax* is the most widespread.^2^ Duffy negative individuals were thought to be resistant to *P. vivax*. However, a growing number of *P. vivax* cases reported throughout Africa where Duffy-negative individuals predominate,^3^ demonstrated that *P. vivax* can infect Duffy-negative individuals^4,5^ and could potentially spread and transmit across populations.^5,6^ Due to epidemiological and ethnic differences, the prevalence of *P. vivax* in Duffy-negative individuals varies across Africa.^4^ Considering *P*. *vivax* can infect and adapt to Duffy-negative individuals, it is possible that these infections can produce gametocytes leading to transmission.^7^ The extent of transmission may vary by environmental and host factors.^8^

During the *Plasmodium* life cycle, the parasites undergo multiple asexual replicative cycles in the human host, and in each erythrocytic replication cycle, a small portion (L0.1%-5%) of the asexual stages develops into sexual gametocytes. The proportion of infections that carries gametocytes is a proxy for human-to-mosquito transmissibility.^9^ Within the mosquito’s midgut, male and female gametocytes undertake gametogenesis.^10^ After the gametes have fertilized, a zygote is created, which later transforms into a motile ookinete. Under the basal lamina, ookinetes form an oocyst by crossing the midgut epithelium.^10,11^ Numerous thousands of sporozoites develop in the oocyst, and as the oocyst wall ruptures, sporozoites enter the hemolymph and infect the salivary gland. The intricate life cycle of the parasite is then completed when sporozoites are inoculated into another person through mosquito bites.^12^ Gametocytogenesis is influenced by epigenetic, ecological, and heritable factors associated with the parasite.^13^ The occurrence of gametocytogenesis is also influenced by factors associated with human hosts such as immunity status, antimalaria drug treatment and genetic factors.^14,15^

The distribution of *P. vivax* in Duffy-negatives across Ethiopia as well as the parasite stages of these infections remain largely unclear. The presence of gametocytes in symptomatic or asymptomatic individuals can lead to onward transmission in the communities. Knowledge of gametocyte reservoirs allows for prioritizing transmission blocking vaccines against *P. vivax* in Africa.^16,17^ This study aims to 1) compare the distribution of *P. vivax* in Duffy-positive and Duffy-negative across Ethiopia; 2) determine the different stages of *P. vivax* in Duffy-positive and Duffy-negative infections; and 3) examine demographic and clinical features of Duffy-negative *P. vivax* infections. These findings will advance current knowledge of *vivax* malaria distribution and transmission in Africa.

## Material & Methods

### Study sites

A total of 447 febrile patient samples were collected in twenty-seven (27) health facilities from seven major regions of Ethiopia including Afar, Amhara, Benishangul/Gumuz, Gambella, Oromia, Sidama, and Southern Nations Nationalities and People’s Region (SNNPR) (**Figure 1**) from 2020-2021. These seven regions also vary in elevation: Afar is in the northeastern part of the country with altitude 379 m (Lat, Long: 11.568, 41.438),^18^ Amhara is in the north with altitude 1268 m (Lat, Long: 11.66334, 338.821903),^19^ Gambella is in the west bordering Sudan with altitude 447 m (Lat, Long: 8.24999, 34.5833),^20^ Oromia is in the east with altitude 959 m (Lat, Long: 7.98906, 39.38118),^21^ Sidama is in the southeast with altitude 1742 m (Lat, Long: 6.7372,38,4008)^22^ and the SNNPR is in the south with altitude 1200 m (Lat, Long: 6.05862, 36.7273).^23^

**Figure 1.**
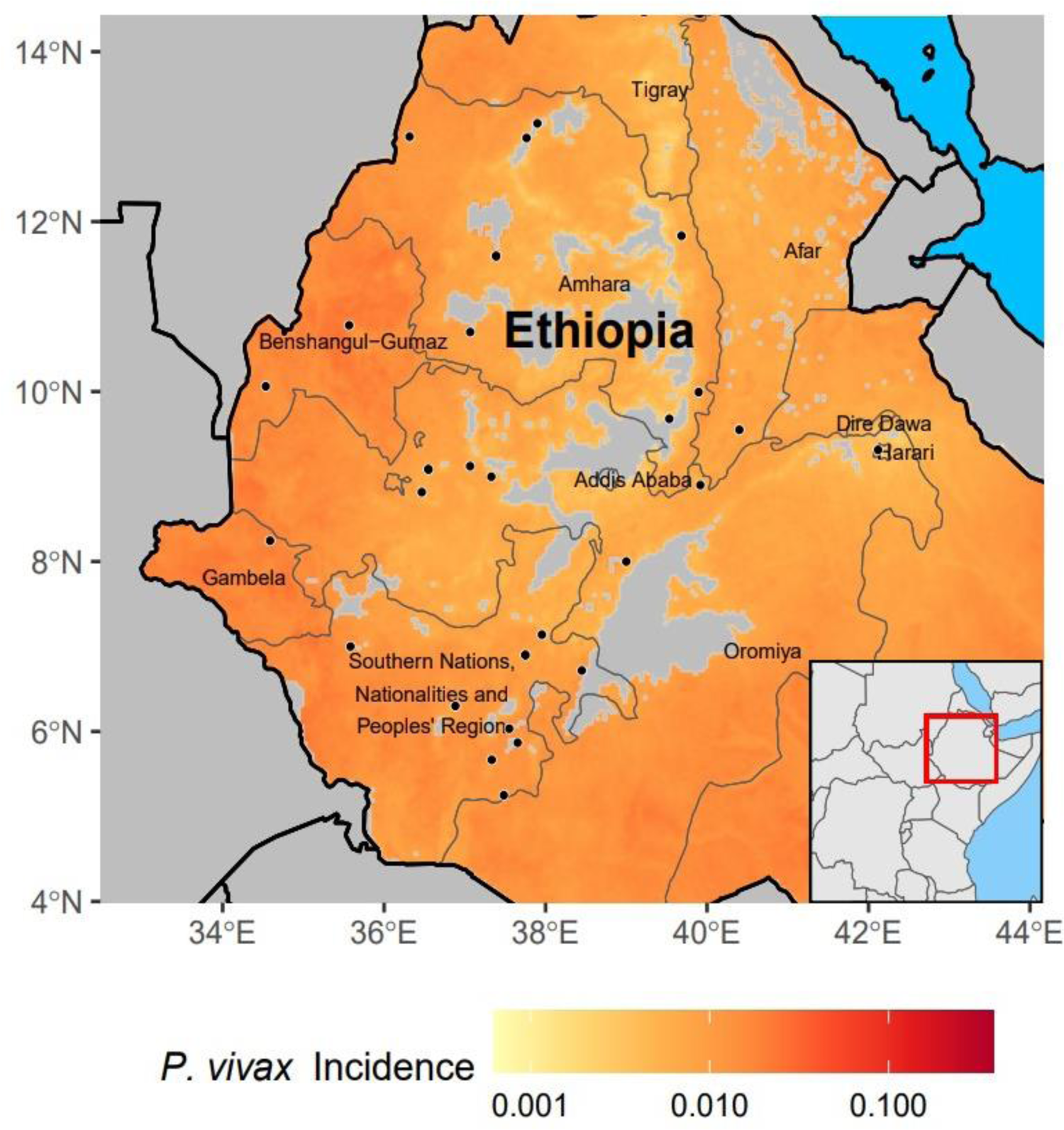
A map showing the study sites in Ethiopia with malaria incidence ranges from high in the western part to low in the eastern part of the country. These sites represent seven major regions including Afar, Amhara, Benishangul/Gumuz, Gambella, Oromia, Sidama, and Southern Nations, Nationalities, and People’s Region (SNNPR) with diverse ethnic groups.

### Blood sample collection and microscopic examination

Finger-prick blood samples were collected from individuals with at least two clinical symptoms, had been suspected for malaria infection and visiting health facilities for malaria diagnosis. All samples were selectively collected from *P. vivax* microscopic confirmed patients.

As soon as possible, the DBS samples were also done at the site of the sample collection for further molecular screening of plasmodium species. The thick and thin blood films were prepared for microscopic screening of *Plasmodium* parasites. Blood smears were stained for 10 minutes with 10% Giemsa staining solution (pH 7.2). The parasite species, the developmental stages of the parasites, and the density of asexual parasites and sexual gametocytes were examined by microscopy. A minimum of 200 microscopic fields were examined at a magnification of 1,000X using oil immersion optics before a slide was declared negative for malaria parasites by the light microscope. Parasitemia/μl of blood was estimated from the thick films as follows: the number of parasites per 200 white blood cells was multiplied by 8,000 (an average white blood cell count/μl) and then divided by 200. Slides were read twice by the primary readers at the site of the study and the secondary readers at EPHI. Discordant results were confirmed by a third slide expert microscope readers. Final species diagnosis was decided by the expert readers. Rapid diagnostic test (RDT) was also conducted for malaria detection.^24,25^

### Molecular screening of *Plasmodium species*

Parasite DNA was isolated from a dried blood spot using the Saponin/Chelex method.^26^ *P. vivax* and *P. falciparum* were detected by the SYBR Green qPCR detection method ^2^ using the published primers (forward: 5′-GAATTTTCTCTTCGGAGTTTATTCTTAGATTGC-3′; reverse: 5′-GCCGCAAGCTCCACGCCTGGTGGTGC-3′) specific to *P. vivax*^27,28^ and *P. falciparum* 18S rRNA (forward: 5′-AGTCATCTTTCGAGGTGACTTTTAGATTGCT-3′; reverse: 5′-GCCGCAAGCTCCACGCCTGGTGGTGC-3′).^29^ Amplification was conducted in a 20 μl reaction mixture containing 2 μl of genomic DNA, 10 μl SYBR Green qPCR Master Mix (Thermo Scientific), and 0.5 uM primer. The reactions were performed in QuantStudio Real-Time PCR Detection System (Thermo Fisher), with an initial denaturation at 95°C for 3 min, followed by 45 cycles at 94°C for 30 sec, 55°C for 30 sec, and 68°C for 1 min with a final 95°C for 10 sec. This was followed by a melting curve step of temperature ranging from 65°C to 95°C with 0.5°C increments to determine the melting temperature of each amplified product. Each assay included positive controls of *P. vivax* Pakchong (MRA-342G) and Nicaragua (MRA-340G) isolates, *P. falciparum* isolates 7G8 (MRA-926) and HB3 (MRA-155), in addition to negative controls including uninfected samples and water. A standard curve was produced from a ten-fold dilution series of the *P. vivax* and *P. falciparum* control plasmid to determine the amplification efficiency of the qPCR. Melting curve analyses were performed for each amplified sample to confirm specific amplifications of the target sequence. The slope of the linear regression of threshold cycle number (*Ct*) versus log10 (gene copy number) was used to calculate amplification efficiency of each plate run based on internal standard controls. For the measure of reproducibility of the threshold cycle number, the mean *Ct* value and standard error was calculated from three independent assays of each sample. A cut-off threshold of 0.02 fluorescence units that robustly represented the threshold cycle at the log-linear phase of the amplification and above the background noise was set to determine *Ct* value for each assay.

Samples yielding *Ct* values higher than 40 (as indicated in the negative controls) were considered negative for *Plasmodium* species. Parasite density in a sample was quantified by converting the *Ct* values into gene copy number (GCN) using the following equation: GCN_sample_ = 2 ^E×(40-^ *^Ct^*^sample)^; where GCN stands for gene copy number, *Ct* for the threshold cycle of the sample, and E for amplification efficiency. The differences in the log-transformed parasite GCN between samples among the study sites were assessed for significance at the level of 0.05.^30,31^

### Duffy blood group genotyping

For all DBS samples, we employed qPCR-based TaqMan assay to examine the point mutation (c.1-67T>C; rs2814778) in the GATA-1 transcription factor binding site of the *DARC* gene. The following primers (forward: 5’-GGCCTGAGGCTTGTGCAGGCAG-3’; reverse: 5’-CATACTCACCCTGTGCAGACAG-3’) and dye-labeled probes (FAM-CCTTGGCTCTTA[*C*]CTTGGAAGCACAGG-BHQ; HEX-CCTTGGCTCTTA[*T*]CTTGGAAGCACAGG-BHQ) were used. Each PCR contained 5μl TaqMan Fast Advanced Master mix (Thermo), 1μl DNA template, and 0.5μl of each primer (10nM), and 0.5μl of each probe (10nM). The reactions were performed with an initial denaturation at 95°C for 2 min, followed by 45 cycles at 95°C for 3 sec and 58°C for 30 sec. A no-template control was used in each assay. The *Fy* genotypes were determined by the allelic discrimination plot based on the fluorescent signal emitted from the allele-specific probes. For *P. vivax* positive samples, a 1,100-bp fragment of the *DARC* gene was further amplified using previously published primers.^3^ Each PCR contained 20μl DreamTaq PCR Mastermix, 1μl DNA template, and 0.5μl each primer. PCR conditions were 94°C for 2-min, followed by 35 cycles of 94°C for 20s, 58°C for 30s, and 68°C for 60s, followed by a 4-min extension. PCR products were purified, and Sanger sequenced. Chromatograms were visually inspected to determine and confirm the *Fy* genotypes based on the TaqMan assays.^31^

### Statistical analyses

SPSS version 21.0 was used for analyzing the socio-demographic information of the participants using descriptive statistics. To test the association between malaria infection and factors including gender, age, ethnicity, and clinical symptoms, bivariate and multivariate logistic regression was performed. The odds ratio and associated 95% confidence interval (CI) were computed to assess the strength of association. *P*-values under 0.05 were considered as significant.

### Ethics statement

Scientific and ethical clearance was obtained from the institutional scientific and ethical review boards of Ethiopian Public Health Institute, Ethiopia and Drexel University, USA. Written informed consent/assent for study participation was obtained from all participants of the study and parents/guardians (for minors under 18 years old).

## Results

### Socio-demographic characteristics of study participants

For the 447 study participants, the age ranged from 6 months to 70 years old. The mean age was 20.87; and 269 (60.1%) of the respondents were males and 171 (38.2%) were females. The greatest proportion of the samples were from SNNPR (34.8%) followed by Oromia (31.3%) and Amhara (23.9%). Afar (1.1%) and Sidama (0.4%) had the smallest sample size (**Table 1**).

**Table 1.** Socio-demographic characteristics of the study participants in seven regional states across Ethiopia.

| Characteristics | Number of participants (%) |
| --- | --- |
| **Gender** |  |
| Male | 269 (60.1%) |
| Female | 171 (38.2%) |
| **Age (years old)** |  |
| < 15 | 150 (33.5%) |
| ≥ 16 and < 45 | 273 (61.0%) |
| ≥ 45 | 16 (3.5%) |
| **Region** |  |
| Afar | 5 (1.1%) |
| Amhara | 107 (23.9%) |
| Benishangual/Gumuz | 22 (4.9%) |
| Gambella | 15 (3.3%) |
| Oromia | 140 (31.3%) |
| Sidama | 2 (0.4%) |
| SNNPR | 156 (34.8%) |

### Distribution of the Duffy genotypes and prevalence of gametocyte across Ethiopia

Of the 447 study participants, 422 (94.4%) were confirmed with *Plasmodium* infections. About 72% of the cases were *P. vivax* infection (322 out of 447), 1.8% were *P. falciparum* infection (8 out of 447), and 20.6% were *P. vivax-P. falciparum* mixed infection (92 out of 447). A total of 20 out of the 447 (4.5%) study participants were Duffy-negative. Eleven (11/20) of the Duffy-negatives were infected with *P. vivax*, five (5/20) were infected with both *P. vivax* and *P. falciparum*, and four (4/20) were not infected. Duffy-negative infections by *P. vivax* were observed in different sites across Ethiopia, specifically in the Amhara, Oromia, Benishangul/gumuz and SNNPR regions but not in Afar, Gambella, and Sidama regions. This could be due to the small sample size in these study sites.

The gametocyte prevalence in Duffy-negative individuals was 31.3% (5 out of 16), with one of them detected in a *P. vivax-P. falciparum* mixed infection. This proportion had no significant difference from Duffy-positive samples which showed 37.7% (150 out of 398), with 26 of them detected in *P. vivax-P. falciparum* mixed infections. Gametocyte stages of *P. vivax* infections were mostly found in the SNNPR, 46.2% (70 out of 155) and Amhara, 30.3% (47 out of 155) followed by Oromia, 12.9% (20 out of 155). There was no gametocyte detected in *P. falciparum* infections. Five gametocyte-positive *P. vivax* infections were detected in Duffy-negatives including two from Amhara, two from SNNPR, and one from Oromia regions (**Table 2**).

**Table 2.** Distribution of the Duffy genotypes among *Plasmodium* species infections, mixed (*Pf* and *Pv*) infections, and gametocyte carriers across Ethiopia.

| Region | Total<br>samples | Duffy-positives |  |  |  |  | Duffy-negatives |  |  |  |  |
| --- | --- | --- | --- | --- | --- | --- | --- | --- | --- | --- | --- |
|  |  | *Pv* | *Pf* | Mixed<br>*Pv-Pf* | Malaria-<br>negative | *Pv* with<br>gametocytes | *Pv* | *Pf* | Mixed<br>*Pv-Pf* | Malaria-<br>negative | *Pv* with<br>gametocytes |
| Afar | 5 | 0 | 0 | 5 | 0 | 0 | 0 | 0 | 0 | 0 | 0 |
| Amhara | 107 | 65 | 4 | 18 | 15 | 45 | 5 | 0 | 0 | 0 | 2 |
| Benishangul/Gumuz | 22 | 6 | 0 | 13 | 0 | 5 | 2 | 0 | 0 | 1 | 0 |
| Gambella | 15 | 7 | 0 | 8 | 0 | 11 | 0 | 0 | 0 | 0 | 0 |
| Oromia | 140 | 116 | 3 | 13 | 1 | 19 | 3 | 0 | 2 | 3 | 1 |
| Sidama | 2 | 2 | 0 | 0 | 0 | 2 | 0 | 0 | 0 | 0 | 0 |
| SNNPR | 156 | 120 | 0 | 26 | 6 | 68 | 1 | 0 | 3 | 0 | 2 |
| Total | 447 | 315 | 7 | 83 | 22 | 150 | 11 | 0 | 5 | 4 | 5 |

### Asexual parasitemia and parasite stage comparisons

No significant difference was detected in parasitemia among the *P. vivax* samples collected from southwestern, southern, and eastern regions of Ethiopia, except for samples in Amhara, which is in the northwest. While previous studies indicated that parasitemia in Duffy-negative individuals are expected to be low, our data showed that *P. vivax* parasitemia in Duffy negatives widely vary among infections, with relatively low parasitemia observed in Oromia and SNNPR regions (Figure 2).

**Figure 2.**
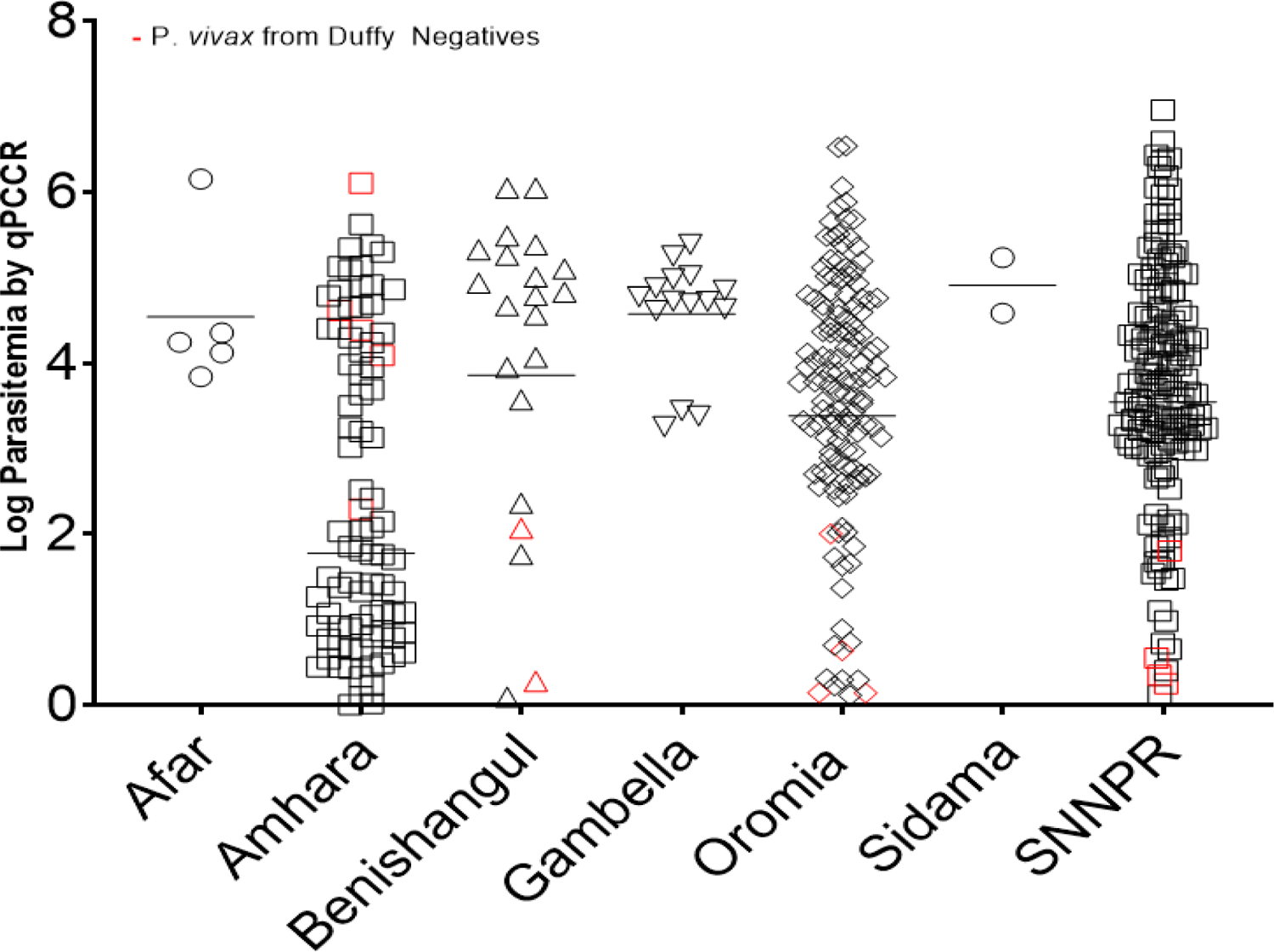
Asexual parasitemia comparison by qPCR among Duffy-positive and Duffy-negative malaria patients in different major regions of Ethiopia.

Most of the infections had mixed parasite stages and the proportion of parasite stages vary among regions. In SNNPR, 71 out of 140 (50.7%) *P. vivax* samples had trophozoites, 66 (47.1%) with mixed trophozoite, schizont, and gametocyte stages, and three (2.1%) with gametocytes only. In Oromia, 99 of the 118 (83.9%) *P. vivax* samples had trophozoites, followed by 16 (13.6%) with mixed trophozoite, schizont, and gametocyte stages, and three (2.5%) with gametocytes only. In Amhara, similar proportion was observed where 55 of 107 (51.4%) *P. vivax* samples had mixed trophozoite, schizont, and gametocyte stages, followed by 45 (42.1%) with trophozoites, and three (2.8%) with gametocytes. In Benshangul/gumuz, 17 of 22 (77.3%) *P. vivax* samples had trophozoites and five (22.7%) had mixed trophozoite, schizont, and gametocytes. In Gambella, 11/15 (73.3%) *P. vivax* samples had trophozoites with gametocyte mixed and gametocyte only. In Afar, all five mixed *P. vivax* and *P. falciparum* samples were detected trophozoites. In Sidama, the two *P. vivax* samples had mixed trophozoite, schizont, and gametocyte stages. Overall, almost all samples had trophozoites and mixed stages across study sites (Figure 3).

**Figure 3.**
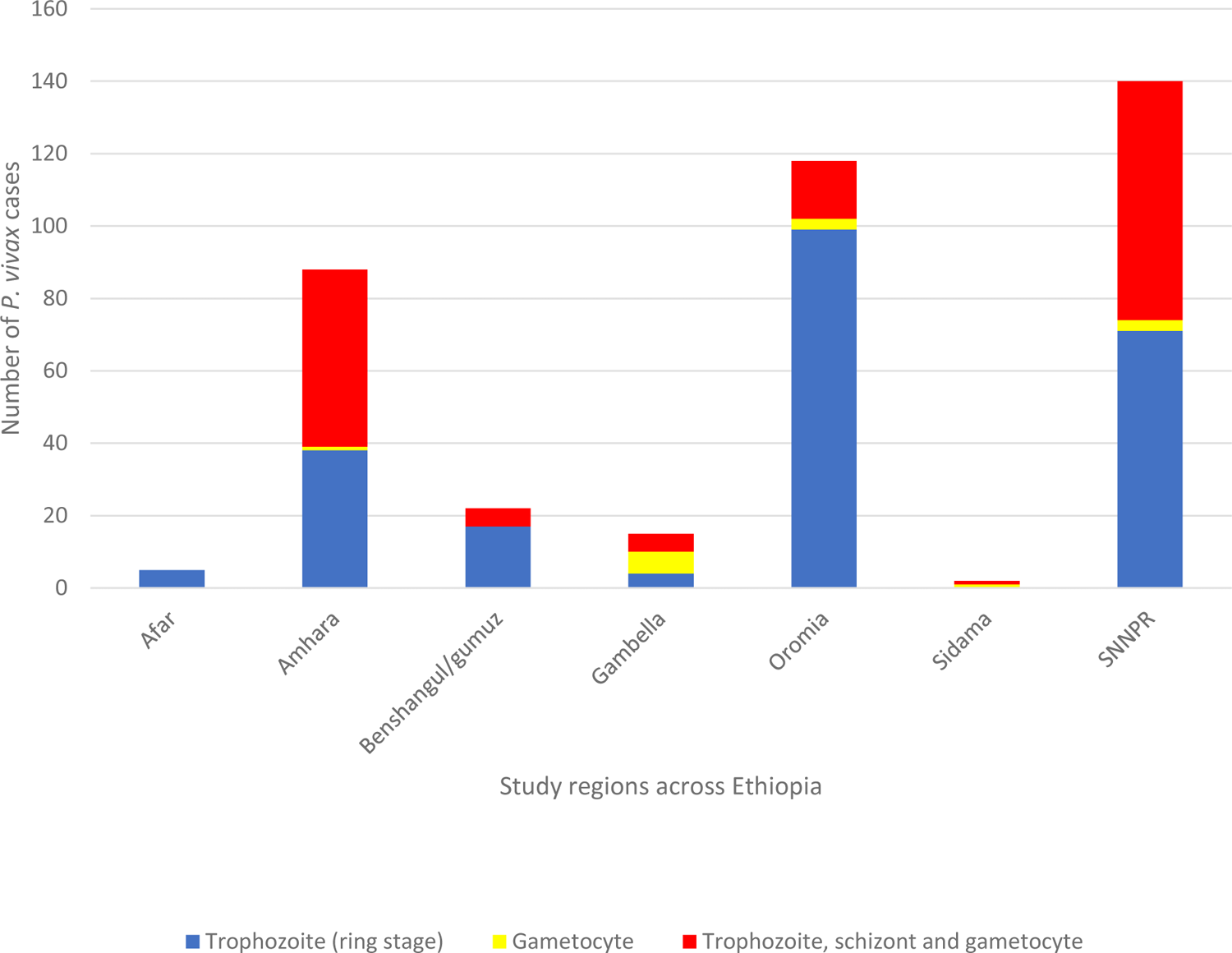
Comparison of parasite stages of *P. vivax*-infected samples by microscopic examination across broad regions of Ethiopia.

The gametocyte count of *P. vivax* species was done with a light microscope against 200 WBCs and calculated using an average WBC count value per μl of blood. The highest gametocyte count was 2856 gametocyte/μl, detected in homozygous Duffy-negative individual and the lowest gametocyte count was 15 gametocytes/μl, detected in heterozygous Duffy-positive individual. The average number of *P. vivax* gametocyte stages among all samples detected with gametocyte stages and all Duffy blood group status was 449 gametocytes/μl of blood, of which the average gametocyte counts of homozygous Duffy negatives (CC), heterozygous Duffy positives (TC) and homozygous Duffy positives (TT) were 1060 gametocytes/μl, 425 gametocytes/μl and 395 gametocytes/μl, respectively (Figure 4).

**Figure 4.**
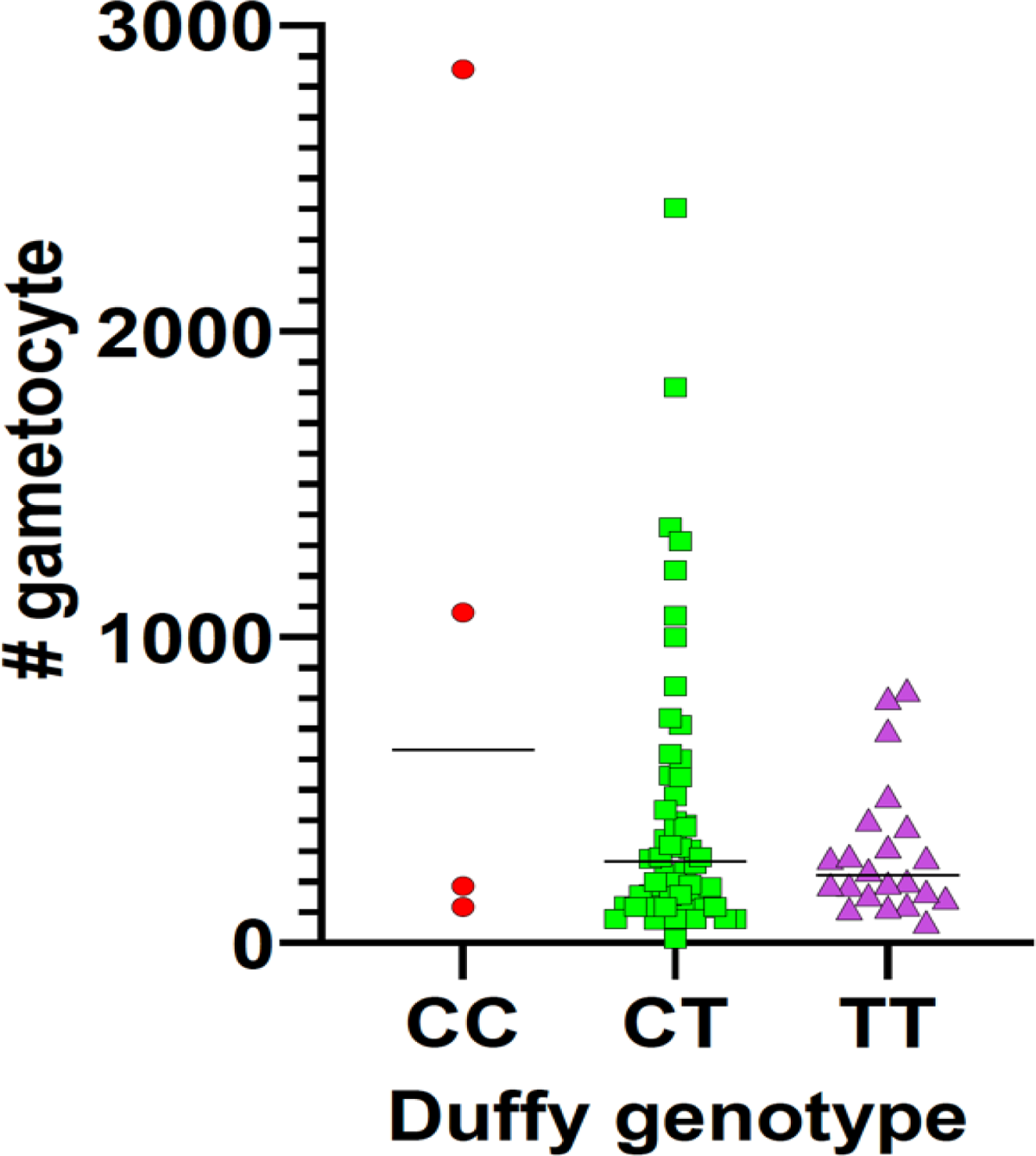
The gametocyte counts among homozygous Duffy-negatives (CC), heterozygous Duffy-positives (TC) and homozygous Duffy-positives (TT) isolates of *P. vivax* species.

### Duffy blood group and other factors associated with *Plasmodium* infections

The bivariate analysis was done to show the association of *P. vivax* infection with independent factors. The prevalence of *P. vivax* infections in Duffy positive individuals were about four times more likely than the Duffy negatives (OR = 4.6, 95% CI 1.4, 14.96, *p* = 0.011).

*P. vivax* infection was not significantly different among males and females. Although the prevalence of *P. vivax* infection was recorded in all age groups, a relatively higher in the age group < 15 years old, three times more likely than the age group > 45 years old (OR = 2.9, 95% CI 0.45-1.98, *p* = 0.98). The *Plasmodium* infections were significantly different among various clinical symptoms of the study participants. The odds of infection among patients with headache was three times more likely than without headache (OR = 3.0, 95% CI 0.81-11.14, *p* = 0.09), patients who had sweating is three times more likely than those who did not have sweating (OR = 3.5, 95% CI 1.7-7.44, *p* = 0.0009) and among patients with chills were over two times more likely than without chills (OR = 2.5, 95% CI 1.05-4.8, *p* = 0.037). No significant difference was found in malaria symptoms such as fever, muscle and joint pain, nausea, and vomiting between plasmodium infected and non-infected individuals (*p* > 0.05) (**Table 3**).

**Table 3.** Results of bivariate odds ratio to determine main predictors of *Plasmodium* infections across Ethiopia. Asterisk represents significance at level of 0.05.

| Parameters |  | Infection rate by 18s qPCR |  |  |  |
| --- | --- | --- | --- | --- | --- |
|  |  | Total<br>samples | Infection | Non-<br>infection | Odds ratio (95% CI) |
| **Duffy**<br>**status** | Duffy+ | 427 | 405 | 22 | 4.6 (1.4, 14.96), $P= 0.011^*$ |
|  | Duffy- | 20 | 16 | 4 | 1 |
| **Gender** | Female | 171 | 159 | 12 | 1 |
| | Male | 269 | 249 | 20 | 0.94 (0.45, 1.98) $P= 0.87$ |
| **Age (years**<br>**old)** | $\leq 15$ | 150 | 143 | 7 | 2.9 (0.55, 15.4) $P= 0.21$ |
| | $> 15$ and $< 45$ | 273 | 250 | 23 | 1.55 (0.33, 7.26) $P= 0.58$ |
| | $\geq 45$ | 16 | 14 | 2 | 1 |
| **Symptoms** |  |  |  |  |  |
| **Fever** | Yes | 398 | 369 | 29 | 0.59 (0.03, 10.43) $P= 0.72$ |
|  | No | 10 | 10 | 0 | 1 |
| **Headache** | Yes | 391 | 365 | 26 | 3.0 (0.81-11.14) $P=0.099$ |
|  | No | 17 | 14 | 3 | 1 |
| **Fatigue** | Yes | 257 | 238 | 19 | 0.89 (0.4, 1.96) $P=0.77$ |
|  | No | 151 | 141 | 10 | 1 |
| **Muscle and**<br>**Joint Pain** | Yes | 257 | 240 | 17 | 1.12 (0.52, 2.41) $P= 0.77$ |
|  | No | 163 | 151 | 12 | 1 |
| **Chills** | Yes | 258 | 245 | 13 | 2.25 (1.05, 4.8) $P=0.037^*$ |
|  | No | 150 | 134 | 16 | 1 |
| **Sweating** | Yes | 218 | 208 | 10 | 3.5 (1.7, 7.44) *P*= 0.0009* |
|  | No | 200 | 171 | 29 | 1 |
| **Nausea and Vomiting** | Yes | 177 | 163 | 14 | 0.81 (0.38, 1.73) *P*=0.59 |
|  | No | 230 | 215 | 15 | 1 |

## Discussion

In Sub-Saharan Africa where Duffy-negative individuals are predominant, *vivax* malaria has been reported but whether these infections can transmit among individuals is poorly documented. This study indicates that *P. vivax* infections in Duffy-negative individuals distribute across broad regions of Ethiopia. In our study, the prevalence of *P. vivax* among Duffy-negatives was 3.8% (16/421). The result is in line with previous studies in the country revealed the prevalence of *P. vivax* among Duffy-negatives 2.9%^32^ and 4.4%,^33^ on the contrary our result is lower than the result found in Sudan (17.9).^34^ In the general populations, Duffy negativity varies from 20–36% in East Africa to 84% in Southern Africa.^4^ Compared to Duffy-positive infections, the average parasite density is much lower in Duffy-negative infections.^4^ Nonetheless, a few Duffy-negative *P. vivax* infections in Amhara were detected with relatively high parasitemia, suggestive of certain *P. vivax* strains can invade Duffy-negative erythrocytes efficiently. The exact mechanisms of Duffy-negative erythrocyte invasion by *P. vivax* are still unclear and merit further investigation. For instance, *P. vivax* glycosylphosphatidylinositol-anchored micronemal antigen (*Pv*GAMA) and merozoite surface protein-1 paralog (*Pv*MSP1P) have been recently shown to bind to both Duffy-positive and negative red blood cells, suggesting their possible involvement in Duffy-independent invasion pathway.^35–37^ The reticulocyte binding protein (*Pv*RBP2b) of *P. vivax* has been shown to bind to transferrin receptor 1 (TfR1) to invade Duffy-positive RBCs and thus, present alternative pathways for Duffy-negative erythrocyte invasion.^5,38^ Such findings are critical to the development of blood-stage vaccine against the parasites.^39–41^

Amongst regions, the difference in *P. vivax* gametocyte production in Duffy-positive and Duffy-negative individuals was not significant. However, the mean number of gametocyte count among Duffy-negatives was higher compared to Duffy-positives, 1060 gametocytes/μl and 425 gametocytes/ μl, respectively. This result indicates the dominance of sexual stages of *P. vivax* parasites that contributes for the occurrence of asymptomatic infections in Duffy-negative individuals. The detection of *P. vivax* gametocytes in Duffy-negative infections in Amhara, Oromia, and SNNPR raises concern that these infections not only cause clinical symptoms but can also contribute to transmission, despite its lower prevalence than in Duffy-positive infected individuals. Given Duffy-negative and Duffy-positive individuals co-exist in Ethiopia, the extent of transmission remains uncertain. It is possible that the asexual parasites converted into gametocytes and spread from Duffy-negative to other Duffy-negative or Duffy-positive individuals.^4,32,42^ This finding lends support to earlier study showing that the parasites detected in Duffy-negative and Duffy-positive populations were not genetically different.^4^ Based on computation modeling, Duffy-negatives in Ethiopia can serve as both the source and sink of infections, though transmission is likely more frequent in Duffy-positives.^4,5^

Given that *P. vivax* has been widely reported in West and Central Africa where > 90% of the populations are Duffy-negatives, these infections can certainly serve as reservoirs for transmission both at the local and regional level.^5,33^ Due to previously being exposed, the host may have acquired immunity against symptomatic blood-stage parasitemia; however, due to the early gametocyte development of *P. vivax*, long lasting sub-clinical illnesses may still contribute to continuous transmission.^43,44^ In this study, almost all gametocytes detected among the mixed infections were *P. vivax*. This result supports the notion that the development of *P. vivax* gametocytes is much faster than the *P. falciparum* ones at the onset of symptoms in febrile malaria patients, and that *P. falciparum* gametocytes are seldomly detected in routine microscopic examination of febrile malaria patients.^45^

Most *P. vivax* infections in Amhara, Gambella, Sidama, and SNNPR had mixed parasite stages including gametocytes, whereas in Oromia and SNNPR, trophozoites were prominent in most samples. This variation in the parasite developmental stages could be associated with environmental, host and parasite factors among different study districts. The epidemiology of malaria within each district may also be a determining factor. For instance, while the general proportion of *P. falciparum* and *P. vivax* in Ethiopia is 60% and 40%, respectively, considerable regional difference exists.^46^

Warmer temperatures and higher rainfall/humidity in lowland than highland areas may allow parasites to develop faster and greater production of gametocytes, which result in majority mixed stages among *P. vivax* infections and enhance transmission. This might be supported by the ability of the parasite to develop the asexual stage into gametocytes faster, within 48 hours after generation of the first merozoites in the blood. The *P. vivax* infected and swollen RBCs due to gametocyte development are flexible and can pass splenic filtration helps to stay all stages together in the peripheral blood.^13^ The unique biological features and genetic variability of the *P. vivax* parasites certainly present a challenge in eradicating malaria in Ethiopia.^47,48^

For all *P. vivax* confirmed infections, typical symptoms were fever as well as headache and fatigue. Other symptoms including muscle and joint pain, chills, sweating, and vomiting vary by individuals across the seven study regions. Interestingly, our analyses revealed that *P. vivax* cases were more likely to occur in individuals aged ≤ 15 years old followed by < 45 and > 15 years old age group. Such demographic pattern could be explained as the host immunity in old individuals might be higher compared to the younger and children. The mosquito vector feeding time and behavior (outdoor or indoor resting/biting), the form of occupation (outdoor or indoor), environment (rural or remote populations), and economic status (poverty) also have contribution.^49^ These factors are critical when identifying disease trends or at-risk population.^5^

To conclude, the prevalence of Duffy-negative individuals among *P. vivax* malaria patients varies across Ethiopia. This study confirms that Duffy-negativity does not completely protect against *P. vivax* infection and these infections are frequently associated with low parasitemia, which may represent hidden reservoirs that can contribute to transmission.

Understanding *P. vivax* transmission biology and gametocyte function via infectivity studies and *in vitro* assays especially in Duffy-negative populations would enhance the treatment and control strategies of *vivax* malaria in Africa. Further study is needed to quantify *Pvs*25 transcripts by qRT-PCR for gametocyte density in Duffy-negative infected samples and to expand sample size that will allow fair comparisons of gametocyte carriage between Duffy-positive and Duffy-negative infections. A deeper comprehension of the association between Duffy-negativity and the invasion processes of *P. vivax* would aid the development of *P. vivax* specific eradication tactics, including substitute antimalarial immunizations other than a Duffy-binding protein-based vaccine.

## Data Availability

All data produced in the present study are available upon reasonable request made to the corresponding author, as per institutional and national legal norms and procedures.

## Acknowledgements

We thank the laboratory staffs at each sample collection health facilities for assisting sample collection and preliminary laboratory work; Adama malaria center expert microscopists for microscopic examinations; and all study participants for their willingness to provide blood sample and information. We also thank Alfred Hubbard for creating the map for Figure 1.

## Authors’ contributions

TTS, EL, SM and EL Conceptualization and designed the study; TTS, MTN, and AA collected sample and perform preliminary laboratory tests; EL, BRA and EL performed molecular laboratory test, analysis, and interpretation; TTS, EL, BRA, JP and EL wrote and reviewed the paper. All authors read and approved the final manuscript.

## Authors’ addresses

Ebony Little, Department of Biological Sciences, University of North Carolina at Charlotte, North Carolina, USA. Tassew T. Shenkutie, Beka R. Abagero, and Eugenia Lo, Department of Microbiology and Immunology, Drexel University, College of Medicine, Philadelphia, PA, USA, and. Meshesha Tsigie Negash, Abnet Abebe, and Sindew Mekasha, Ethiopian Public Health Institute, Addis Ababa, Ethiopia, and. Jean Popovici, Institute Pasteur in Cambodia, Phnom Penh, Cambodia.

